# A Radiologic Masquerade: Camrelizumab-Associated Breast Lesions That Mimic Progression

**DOI:** 10.64898/2026.05.30.26353749

**Authors:** Yangling Hu, Yanhong Shui, Wei Li, Jialu Liang, Yutong Song, Mengyi Wang, Fan Zhang, Min Zhang, Huan Wang, Lin Ji, Meizhi Li, Chaoyang Wang, Nan Shao, Xiaying Kuang, Shaofu He, Xiaoling Zhang

**Affiliations:** First Affiliated Hospital of Sun Yat-sen University, Guangzhou, China, Department of Radiology, 510080; First Affiliated Hospital of Sun Yat-sen University, Guangzhou, China, Breast Disease Center, 510080; Department of Endocrinology, Armed Police Corps Hospital of Guangdong Province, Guangzhou, China; Department of Radiology, Huadu District People’s Hospital of Guangzhou, China

**Keywords:** Camrelizumab-associated breast lesions (CABLs), Immune-related adverse events (irAEs), Reactive cutaneous capillary endothelial proliferation (RCCEP), Pseudoprogression (PsP)

## Abstract

**Background:** Immune-related adverse events (irAEs) involving the breast remain rarely reported.

**Purpose:** To characterize clinical and imaging features of camrelizumab-associated breast lesions (CABLs).

**Materials and Methods:** This retrospective dual-cohort study (October 2019 to February 2026) included 196 female patients. Cohort A comprised 180 non-breast cancer patients; Cohort B comprised 16 breast cancer patients receiving neoadjuvant camrelizumab. Baseline characteristics, treatment response, and CT/MRI features were compared between CABL-positive and CABL-negative groups using Mann-Whitney U and chi-square tests.

**Results:** CABLs developed in 34.4% (62/180) of Cohort A and 93.8% (15/16) of Cohort B. CABL□positive patients were younger (median 50.5 vs 54.5 years; P = 0.006) and more often premenopausal (46.8% vs 26.3%; P = 0.009). The objective response rate was relatively high among patients with positive lesions; in Group A, the disease progression rate was lower in the CABL-positive group than in the CABL-negative group (3.2% vs 17.8%), whilst in Group B, the pathological complete response rate was as high as 53.3% (8/15). On CT/MRI, CABLs were predominantly multiple (62.5%), with well□defined margins and unrestricted diffusion. The predominant time□intensity curve (TIC) pattern was washout (46.7%). Median time to onset was 2-3 cycles (the second MRI scan); most lesions disappeared (40.3%) and shrank (46.8%) during follow□up. ADC values of lesions were significantly higher than those of primary tumors (1.847±0.284 vs 0.976±0.055 ×10□³ mm²/s; P < 0.001). Histopathology of four lesions revealed lymphocytic infiltration and fibrosis without malignancy.

**Conclusion:** CABLs are benign reactive changes driven by multiple factors. Their recognition prevents misinterpretation as disease progression, thereby avoiding unnecessary treatment discontinuation or biopsy.

**Highlights:** *What is already known on this topic:* Although irAEs have been described in nearly every organ system, involvement of the breast—whether as a primary or secondary site—has been rarely reported.

*What this study adds:* Camrelizumab-associated breast lesions (CABLs) are predominantly benign reactive changes (irAEs), not disease progression, and patients with these lesions show higher objective response rates, suggesting they may serve as a radiological biomarker for better treatment outcomes.

*How this study might affect research, practice or policy:* For CABLs, a “watch and wait” strategy with imaging follow-up is recommended unless atypical features arise.

## Introduction

Immune checkpoint inhibitors (ICIs) have become a cornerstone of cancer therapy. By targeting programmed death-ligand 1 (PD-L1), programmed death-1 (PD-1), or cytotoxic T-lymphocyte-associated antigen 4 (CTLA-4), ICIs disrupt immune evasion mechanisms and enhance T-cell-mediated tumor elimination^[1, 2]^. These inhibitors often induce durable responses that persist after treatment cessation^[3]^. However, as survival improves, immune-related adverse events (irAEs)—resulting from excessive immune activation and loss of self-tolerance—have emerged as a growing challenge. Although irAEs have been described in nearly every organ system, involvement of the breast—whether as a primary or secondary site—has been rarely reported.

Camrelizumab, a PD-1 inhibitor approved for multiple malignancies, has a distinctive irAE profile, most notably reactive cutaneous capillary endothelial proliferation (RCCEP), a low-grade adverse event characterized by proliferation of capillaries and endothelial cells, attributed to off-target activation of VEGF receptor 2 ^[4]^. Camrelizumab is not a first-line immunotherapeutic agent for breast cancer, whether it can induce similar benign proliferative changes in breast tissue remains unknown.

A phase II trial at our center enrolled 16 patients with HRR-mutated, HER2-negative breast cancer who received neoadjuvant camrelizumab combined with nab-paclitaxel and fluzoparib. Among the patients in this trial, we observed that a majority developed new enhancing breast lesions on early follow-up MRI. Initially indistinguishable from residual disease or progression, these lesions subsequently shrank or disappeared on follow-up imaging with continued treatment. To further investigate the potential association between camrelizumab and newly enhanced breast lesions, we retrospectively reviewed contrast-enhanced chest CT imaging of female patients received at least one dose of camrelizumab between October 2019 and December 2025 for the malignant tumors in any location of the body. As expected, newly developed breast lesion was identified in some of the the patients. No new breast lesions were observed in 78 female patients with malignancies who received other PD-1 inhibitor (such as pembrolizumab, sintilimab) on contrast-enhanced CT imaging. We therefore attributed these breast lesions to camrelizumab and refer to them as camrelizumab-associated breast lesions (CABL).

Misinterpretation of these benign treatment-induced lesions as disease progression could lead to unnecessary treatment discontinuation, invasive biopsies, or even surgical excision. Given the limited documentation of breast irAEs, we analyzed the female patients who received camrelizumab for malignancy. Two cohort was constructed, including Cohort A and Cohort B. Cohort A included all female patients with active malignancies other than breast cancer. Cohort B consisted of all the patient with breast cancer. The objective of this study is to characterize the clinical, radiological, and prognostic features of the patients who developed CABL. And to provide practical guidance for differentiating these benign lesions from true primary tumor progression or metastasis, thereby preventing unnecessary interventions and ensuring optimal continuation of immunotherapy.

## MATERIALS AND METHODS

### Study Design and Participants

This dual-cohort retrospective study was conducted at XXX Hospital of XXX University between October 2019 and February 2026. Follow-up data were collected until February 2026 (data cutoff). The inclusion criteria were as follows: (1) female patients with histopathologically confirmed malignancy who received at least one dose of camrelizumab (neoadjuvant or palliative); (2) availability of at least one baseline and two post-treatment contrast-enhanced chest CT (Cohort A) or breast MRI (Cohort B); (3) complete clinical data. Exclusion criteria: (1) prior ICI therapy or thoracic radiotherapy; (2) clinical or pathological breast disease, breast implants; (3) poor image quality; (4) incomplete data.

Initially, 199 patients in Cohort A and 16 patients in Cohort B were screened. To ensure adequate camrelizumab exposure for evaluating CABL development, patients who received only one cycle were excluded (n=19). The remaining 180 patients were enrolled in Cohort A. All 16 patients with breast cancer who received neoadjuvant camrelizumab-based combination therapy were enrolled in Cohort B. **Figure 1** presents the study workflow, cohort composition, and clinical outcomes of their primary malignancies. The study protocol (Approval No. 2022-203) was approved by the Institutional Review Board; informed consent was waived. The study was conducted in accordance with the Declaration of Helsinki.

**Figure 1.**
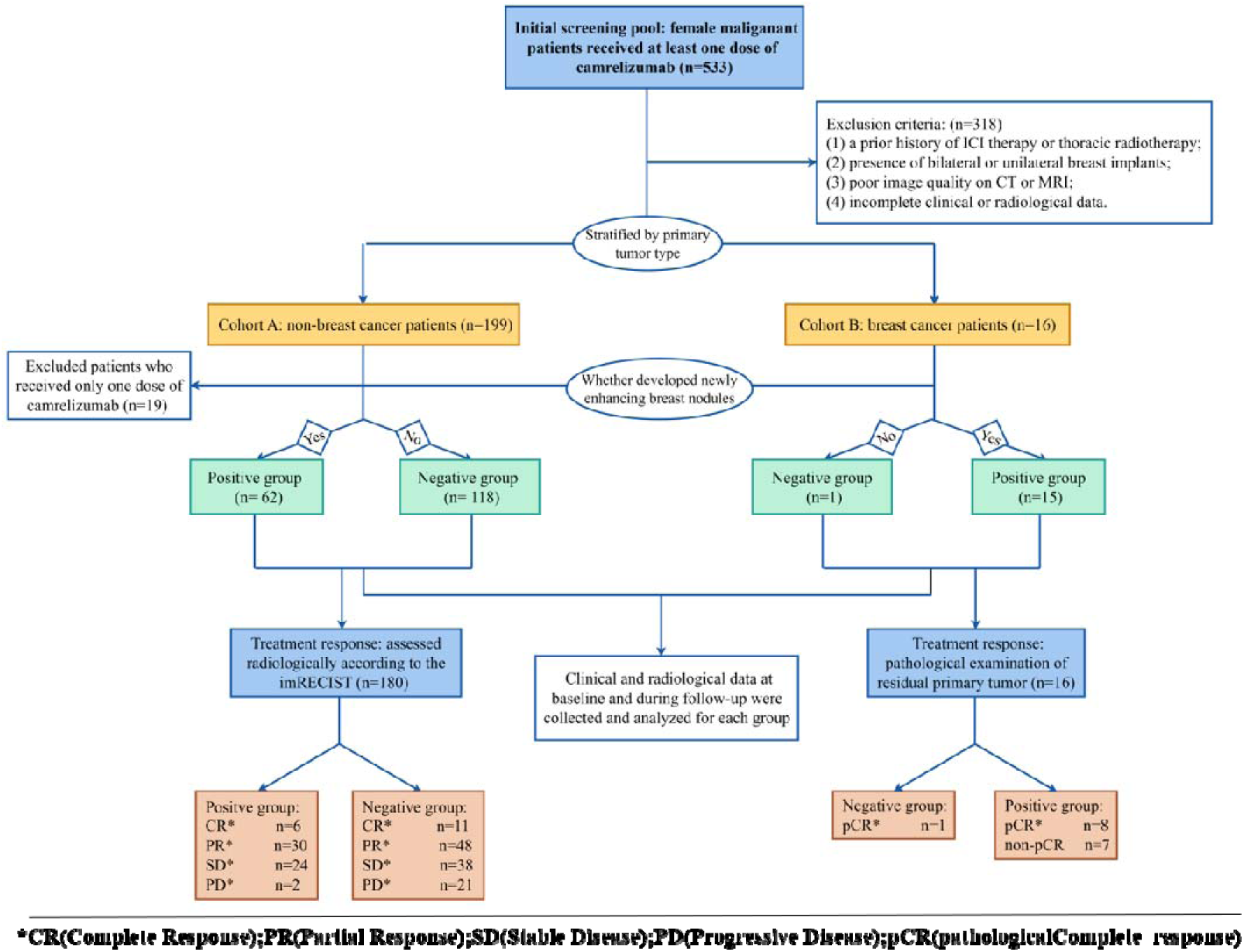
The study workflow.

### Clinical Data Collection and Therapy Efficacy Evaluation

Baseline clinical data were extracted, including age, menstrual status, history of breast disease, primary tumor organ (lung, esophagus, or other), timing of camrelizumab initiation, treatment line (first-line vs. ≥second-line), and number of treatment cycles (categorized as 2, 3-4, ≥5). In longitudinal oncological imaging assessment, the time of emergence of CABL, as well as the timing of their regression or complete disappearance, should be meticulously documented.

In cohort A, most patients received camrelizumab treatment as palliative immunotherapy without surgical intervention. Therapeutic efficacy was assessed using the immune-modified Response Evaluation Criteria in Solid Tumors (imRECIST). Contrast-enhanced whole-body CT scans were conducted every two treatment cycles for response evaluation. Response categories comprised complete response (CR), partial response (PR), stable disease (SD), and progressive disease (PD), with objective response rate (ORR) defined as the proportion of patients attaining CR or PR.

In Cohort B, all patients underwent surgery after neoadjuvant therapy with camrelizumab, enabling pathological assessment with pathologic complete response (pCR) as the primary endpoint. Pathologic complete response was defined as ypT0/Tis ypN0 per the Residual Cancer Burden criteria. In the four patients presenting with CABL, either surgical excision (in 3 patients) or fine-needle aspiration (FNA) (in 1 patient) biopsy was carried out within two weeks following lesion detection, and pathological confirmation was obtained.

### Imaging Acquisition

In Cohort A, contrast-enhanced chest CT was acquired in all patients. All contrast-enhanced chest CT scans parameters were as shown in **Supplementary Table 1.** A total of 80–100 mL of non-ionic iodinated contrast agent (iohexol 300 mgI/mL) was injected via an antecubital vein using a power injector at a flow rate of 2.5-3.0 mL/s, followed by a 20-mL saline flush at the same rate. Contrast-enhanced CT scanning of the chest and whole abdomen was performed at a delay of 65-70 seconds after intravenous contrast administration.

In Cohort B, breast MRI examinations were performed in all 15 patients. The breast MRI was performed on a 3T MRI scanner (Verio, Siemens Healthcare, Erlangen, Germany) equipped with a dedicated 16-channel phased-array breast coil. Transtransal T1WI, T2WI, and DWI were acquired before contrast agent injection. Dynamic contrast-enhanced T1WI was acquired after contrast agent injection. MRI scanning parameters were listed in **Supplementary Table 2**. Breast MRI was acquired before camrelizumab therapy as baseline imaging, and following every two treatment cycles for response evaluation.

### Imaging Analysis

CABL were new lesions detected on both dynamic contrast-enhanced MR sequences (especially during the first two scans) and contrast-enhanced CT imaging, compared with baseline imaging (MRI or CT) obtained prior to Camrelizumab treatment. Importantly, background parenchymal enhancement (BPE) could be unequivocally ruled out.

Radiological features of the CABLs were analyzed on enhanced CT and MRI imaging, according to the Breast Imaging Reporting and Data System (BI-RADS). For mass lesions, the assessed characteristics included size, margin, boundary, enhancement pattern, dynamic contrast-enhanced (DCE) kinetics, and diffusion characteristics. For non-mass lesions, the analysis encompassed distribution pattern and enhancement characteristics. In cases with multiple lesions, the largest lesion was selected for analysis. Longitudinal changes in CABL across multiple imaging time points was also focused on, encompassing alterations in lesion size, lesion count, enhancement patterns, and diffusion characteristics.

Two breast radiologists (5 years’ experience) independently reviewed all contrast-enhanced CT and breast MR images. Disagreements were resolved by a third senior radiologist (15 years’ experience).

### Statistical Analysis

Statistical analyses were performed using SPSS Statistics version 27.0 (IBM Corp, Armonk, NY). Continuous variables were presented as median (interquartile range) or mean ± standard deviation, as appropriate. Categorical variables were presented as frequencies (percentages). Comparisons between CABLs-positive and CABLs-negative groups were performed using the Mann-Whitney U test for continuous variables and the chi-square test (or Fisher’s exact test when expected cell counts <5) for categorical variables. A two-sided P < 0.05 was considered statistically significant.

## Result

### In Cohort A

#### Clinical and pathological features

Baseline clinical characteristics of Cohort A are summarized in **Table 1**. Among the 180 patients in Cohort A, 62 (34.4%) developed CABL during Camrelizumab treatment (CABL-positive group), whereas 118 (65.6%) did not (CABL-negative group). The patients in the positive group were significantly younger than those in the CABL-negative group (median age: 50.5 years [IQR 42.0–57.0] vs. 54.5 years [IQR 49.0–63.0], *P = 0.006*). The proportion of premenopausal patients was larger in the CABL-positive group than in the CABL-negative group (46.8% vs. 26.3%, *P=0.009*). No significant differences were observed between the two groups in the distribution of prior treatment line (*P=0.822*) or the number of treatment cycles (*P=0.364*).

**Table 1.**
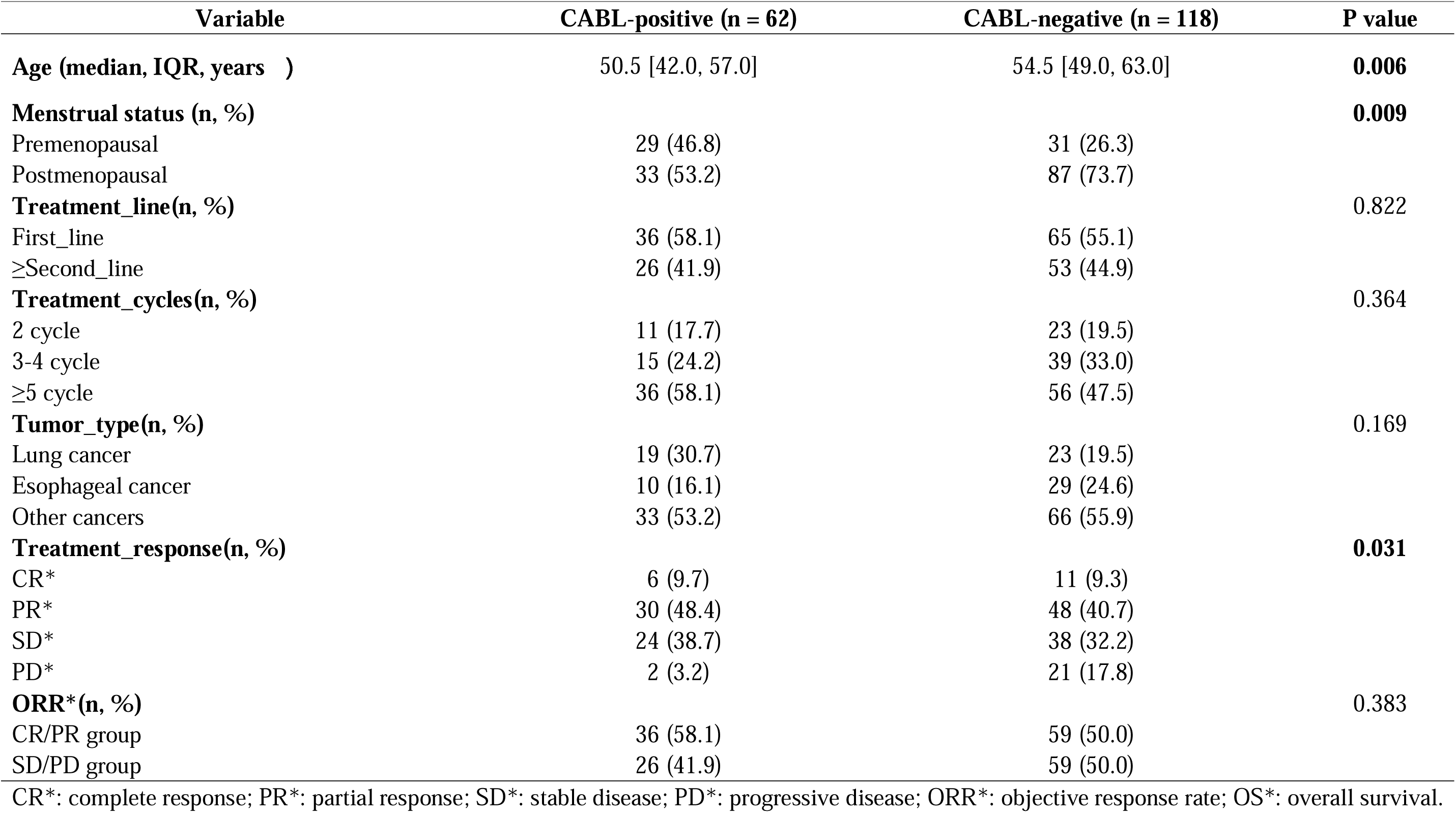
The baseline clinical characteristics and treatment response in Cohort A (n=180)

In Cohort A, there were 42 (23.3%) patients with lung cancer, 39 (21.7%) with esophageal cancer, and 99 (55.0%) with other tumor types (including head and neck cancer, liver cancer, gynecologic tumors, and urologic tumors). In the CABL-positive group of 62 patients, the numbers of patients with lung cancer, esophageal cancer, and other tumors were 19 (30.7%), 10 (16.1%), and 33 (53.2%), respectively. In addition, CABL was found in 19 patients (45.2%) with lung cancer, in 10 patients (25.6%) with esophageal cancer, and in 33 patients (33.3%) with other tumors, respectively, with no statistically significant differences among the three groups (*P = 0.169*) (**Figure 2**). The distribution of response categories differed significantly between the two groups (*P* = 0.031). In particular, the proportions of patients with PD were markedly lower in the CABL-positive group than in the CABL-negative group (3.2% [2/62] vs. 17.8% [21/118]).

**Figure 2.**
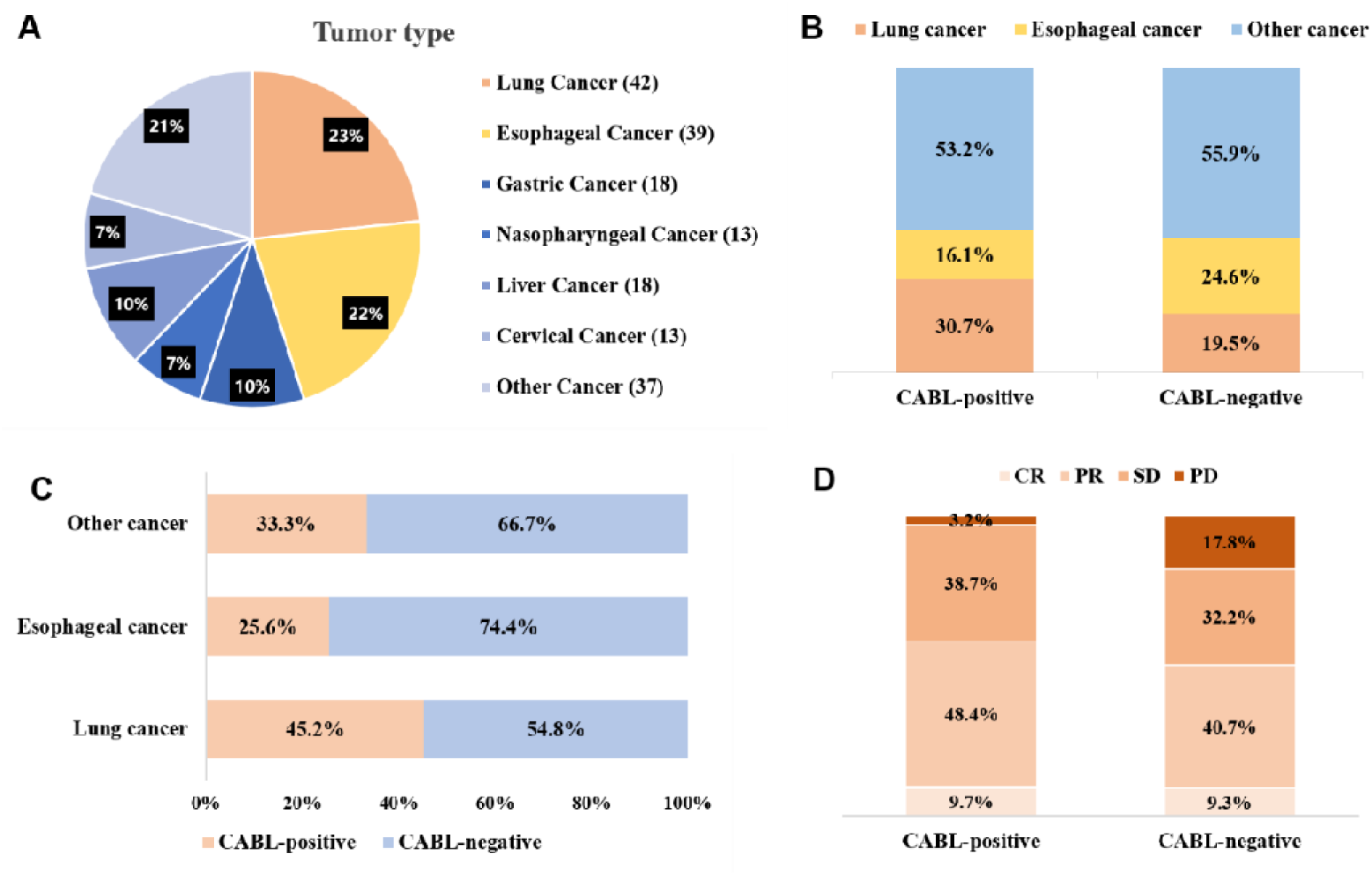
Tumor type distribution and tumor response in Cohort A. **(A)** Distribution of cancer types in Cohort A; (B) The proportions of three major tumor categories (lung cancer, esophageal cancer, and other cancers combined) within CABL-positive and CABL-negative lesion groups; (C) The proportions of CABL-positive patients within the three major tumor categories; (D) The proportions of CR, PR, SD, and PD in the CABL-positive group and the CABL-negative group.

#### Imaging features

The radiological features of CABLs in Cohort A are summarized in **Table 2**. Among the 62 patients in the CABL-positive group, 32 (51.6%) presented with a single new breast lesion, while the remaining 30 (48.4%) had multiple lesions. CABL in bilateral breast was observed in 21 patients (33.9%). Of those with multiple lesions, 26 (41.9%) exhibited involvement of more than one quadrant. Mass-like lesions were found in 43 (69.4%) patients, and non-mass-like lesions in 19 (30.6%) patients. The median diameter of mass-like and non-mass-like lesions was 6 mm (range, 5-15mm) and 9 mm (range, 5-21mm), respectively. Among the mass-like lesions, 40 had regular margins and 3 had irregular margins, the latter posing a risk of misdiagnosis as malignancy. Among the non-mass-like lesions, 10 showed segmental distribution, and 9 showed clustered distribution. Most CABLs (95.2%, 59/62) appeared on follow-up imaging after 2 or 3 treatment cycles. Of these, 48 (77.4%) regressed or disappeared on re-evaluation after 4–5 cycles of treatment. One case showed unchanged CABL during the entire follow-up, while 7 cases were excluded from outcome analysis owing to missing subsequent imaging data.

**Table 2.** CT features of CABLs in Cohort A (n=62)

| **Variable** |  | **CABLs (n=62)** |
| --- | --- | --- |
| **No.of Lesions (n, %)** |  |  |
| single |  | 32 (51.6) |
| multi-focal |  | 30 (48.4) |
| **Distribution (n, %)** |  |  |
| Location |  |  |
|  | unilateral | 41 (66.1) |
|  | bilateral | 21 (33.9) |
| Quadrant |  |  |
|  | single-quadrant | 36 (58.1) |
|  | multi-quadrant | 26 (41.9) |
| **Shape feature (n, %)** |  |  |
| Mass enhancement |  | 43 (69.4) |
|  | regular | 40 (93.0) |
|  | irregular | 3 (7.0) |
| Non-mass enhancement |  | 19 (30.6) |
|  | segmental | 10 (52.6) |
|  | non-segmental | 9 (47.4) |
| **#Dmax\* /mm** |  |  |
| Mass-like |  | 6 [5, 15] |
| Non-mass-like |  | 9 [5, 21] |
| **Onset_time** |  |  |
| cycle 2-3 |  | 59 (95.2) |
| cycle 4-5 |  | 1 (1.6) |
| cycle 6-7 |  | 2 (3.2) |
| **Outcome_final(n, %)** |  |  |
| disappear |  | 25 (40.3) |
| shrink |  | 29 (46.8) |
| unchanged |  | 1 (1.6) |
| missing |  | 7 (11.3) |
| **Time_outcome** |  |  |
| cycle 4-5 |  | 48 (77.4) |
| cycle 6-7 |  | 4 (6.5) |
| ≥ cycle 8 |  | 3 (4.8) |
| missing |  | 7 (11.3) |

#Median [MIN, MAX]; Dmax\*: the maximum diameter.

### In Cohort B

#### Clinical and pathological Features

The clinicopathological features of Cohort B are summarized in **Table 3**. In Cohort B, 16 patients with breast cancer were enrolled. Among them, 15 (93.8%) developed CABL during neoadjuvant camrelizumab therapy, whereas only one patient showed no new breast lesions on serial follow-up imaging. The mean age of the 15 CABL-positive patients was 42.8 years (range, 27-55 years), and 86.7% (13/15) were premenopausal. Histological examination revealed invasive ductal carcinoma in 14 (93.3%) patients, papillary carcinoma in one, and high-grade tumors accounted for 66.7% (10/15). 10 (66.7%) patients had clinical stage III disease. Regarding molecular subtypes, Luminal B was identified in 7 (46.7%) patients, TNBC in 5 (33.3%), and Luminal A in 3 (20.0%). BRCA1 mutations were present in 6 (40.0%) patients, and BRCA2 mutations in 5 (33.3%). Pathologically confirmed positive lymph node metastasis was observed in 12 (80.0%) patients. After camrelizumab-based neoadjuvant combination chemotherapy, 8 (53.3%) patients achieved pCR. All 15 patients underwent breast surgery, breast-conserving surgery (BCS) in 6 (40.0%) patients, modified radical mastectomy (MRM) in 4 (26.7%) patients, and nipple-sparing mastectomy (NSM) in 5 (33.3%) patients.

**Table 3.**
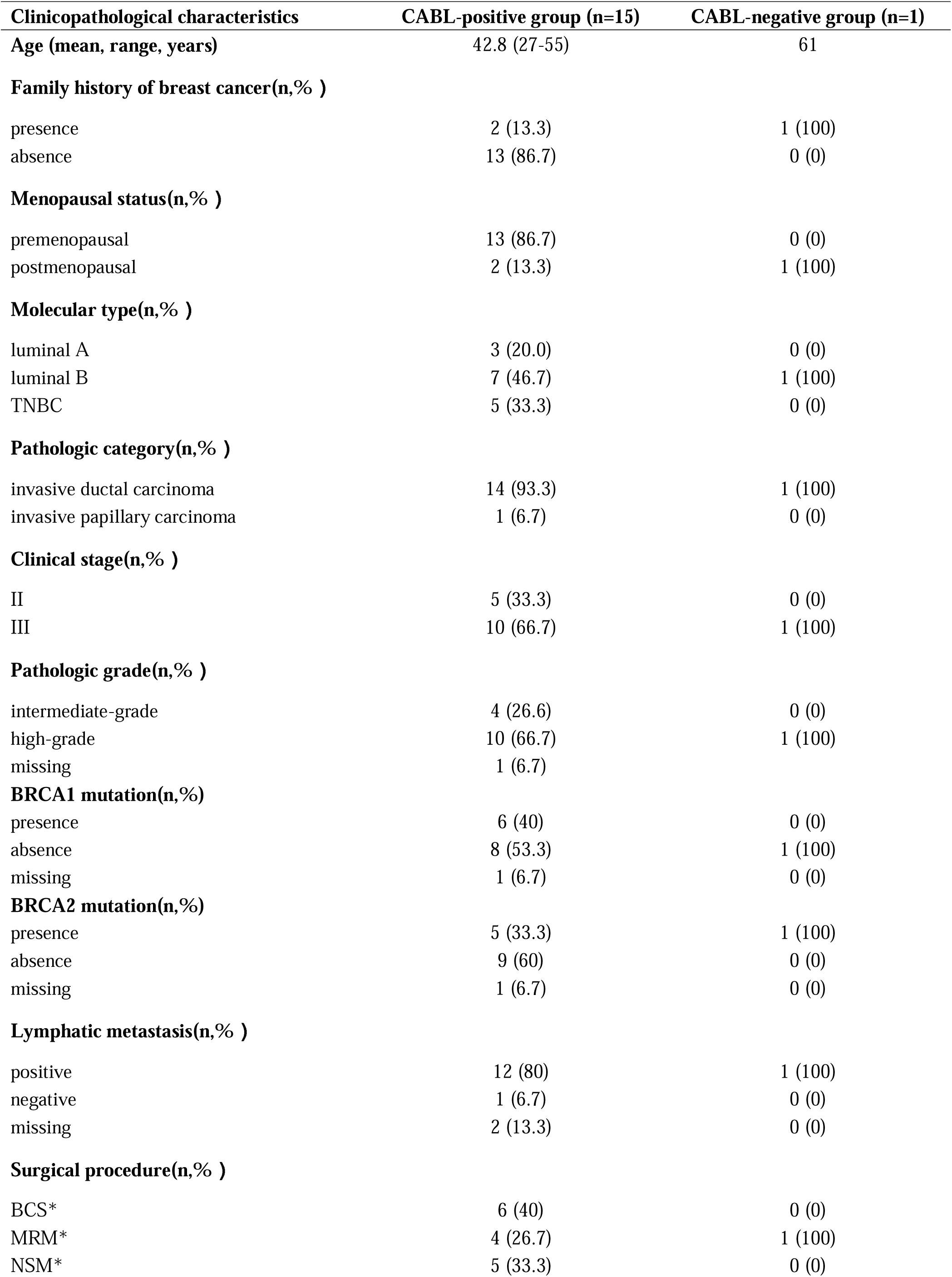
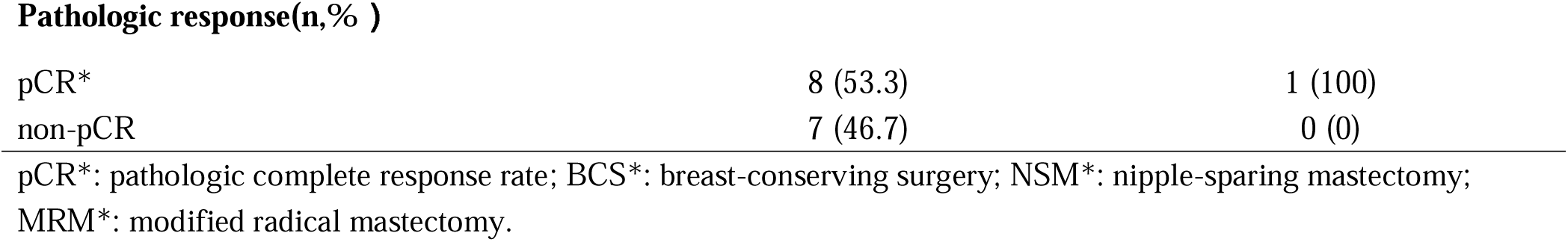
Cohort characteristics and treatment response in Cohort B (n=16)

| **Clinicopathological characteristics** | **CABL-positive group (n=15)** | **CABL-negative group (n=1)** |
| --- | --- | --- |
| **Age (mean, range, years)** | 42.8 (27-55) | 61 |
| **Family history of breast cancer(n,% )** |  |  |
| presence | 2 (13.3) | 1 (100) |
| absence | 13 (86.7) | 0 (0) |
| **Menopausal status(n,% )** |  |  |
| premenopausal | 13 (86.7) | 0 (0) |
| postmenopausal | 2 (13.3) | 1 (100) |
| **Molecular type(n,% )** |  |  |
| luminal A | 3 (20.0) | 0 (0) |
| luminal B | 7 (46.7) | 1 (100) |
| TNBC | 5 (33.3) | 0 (0) |
| **Pathologic category(n,% )** |  |  |
| invasive ductal carcinoma | 14 (93.3) | 1 (100) |
| invasive papillary carcinoma | 1 (6.7) | 0 (0) |
| **Clinical stage(n,% )** |  |  |
| II | 5 (33.3) | 0 (0) |
| III | 10 (66.7) | 1 (100) |
| **Pathologic grade(n,% )** |  |  |
| intermediate-grade | 4 (26.6) | 0 (0) |
| high-grade | 10 (66.7) | 1 (100) |
| missing | 1 (6.7) |  |
| **BRCA1 mutation(n,% )** |  |  |
| presence | 6 (40) | 0 (0) |
| absence | 8 (53.3) | 1 (100) |
| missing | 1 (6.7) | 0 (0) |
| **BRCA2 mutation(n,% )** |  |  |
| presence | 5 (33.3) | 1 (100) |
| absence | 9 (60) | 0 (0) |
| missing | 1 (6.7) | 0 (0) |
| **Lymphatic metastasis(n,% )** |  |  |
| positive | 12 (80) | 1 (100) |
| negative | 1 (6.7) | 0 (0) |
| missing | 2 (13.3) | 0 (0) |
| **Surgical procedure(n,% )** |  |  |
| BCS* | 6 (40) | 0 (0) |
| MRM* | 4 (26.7) | 1 (100) |
| NSM* | 5 (33.3) | 0 (0) |
| **Pathologic response(n,% )** |  |  |
| pCR* | 8 (53.3) | 1 (100) |
| non-pCR | 7 (46.7) | 0 (0) |

pCR\*: pathologic complete response rate; BCS\*: breast-conserving surgery; NSM\*: nipple-sparing mastectomy; MRM\*: modified radical mastectomy.

#### Imaging features

The baseline MRI characteristics of primary tumors in Cohort B are summarized in **Table 4**. For the radiological characteristics of primary tumors in the CABL-positive group, 9 (60.0%) presented multiple lesions, while 6 (40.0%) had a single lesion. 12 (80.0%) patients had minimal to mild BPE (BPE<50%), and moderate to marked BPE (BPE≥50%) was shown in 3 (20.0%) patients. Mass-like lesions were found in 11 (73.3%) patients; of these, 7 had heterogeneous enhancement and 4 had rim enhancement. Non-mass-like enhancement was present in 4 (26.7%) patients, among whom 3 showed segmental distribution, and 1 showed multiregional distribution. 12 (80.0%) lesions exhibited restricted diffusion on DWI and ADC, with a mean ADC value of (0.976±0.055) ×10□³ mm²/s. The time-intensity curve (TIC) was typically a plateau pattern (80.0%, 12/15), while 3 (20.0%) were in a wash-out pattern. The dynamic changes of primary tumors were presented in **Figure 3A and 3C**; the mean diameter of the primary tumors was 40.2±12.0 mm, which decreased to 20.2±14.1 mm at follow-up; the ADC values of most primary tumors were progressively increased during treatment (details in **Supplementary Table 3**).

**Figure 3.**
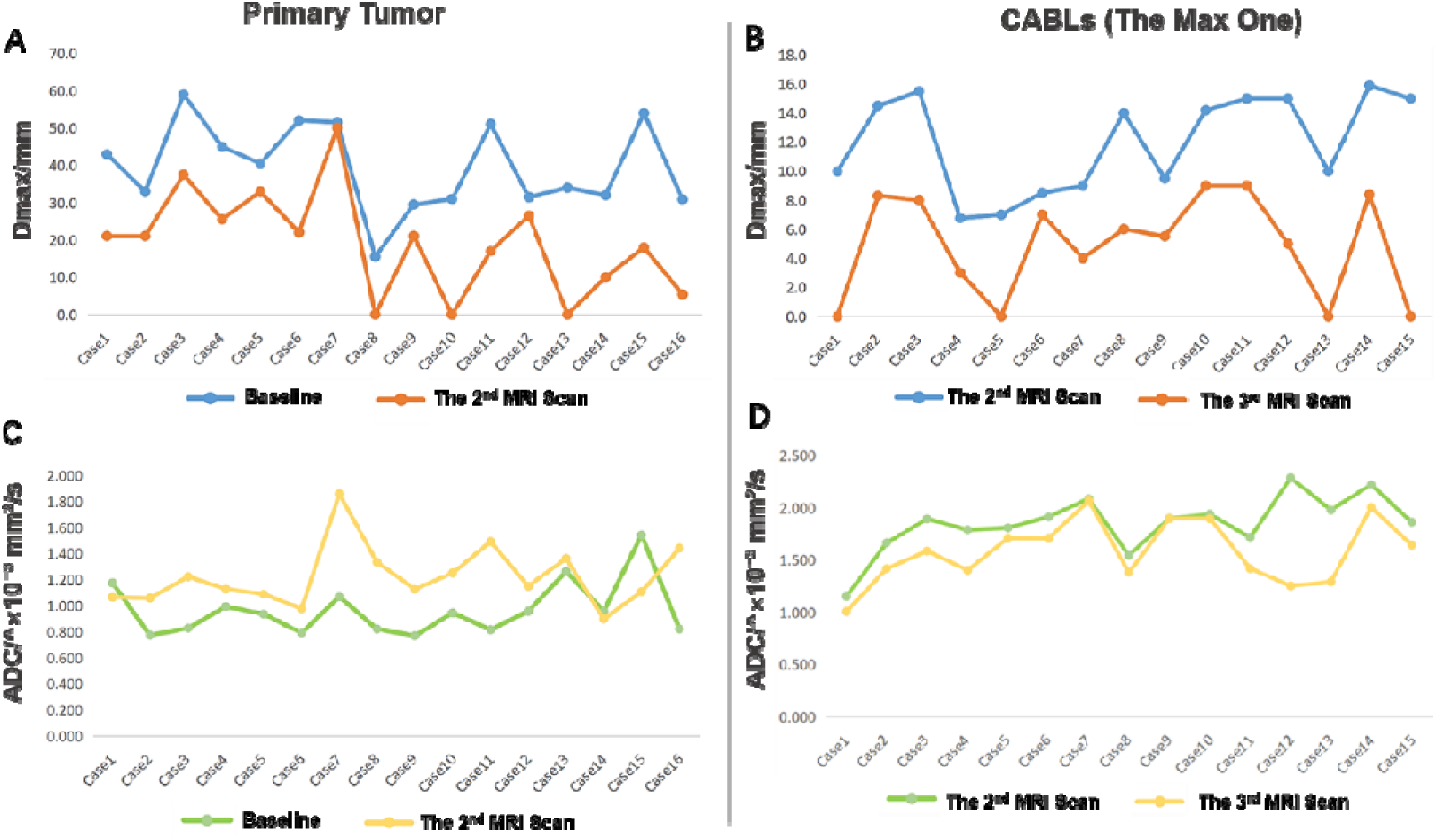
Trends observed on follow-up MRI scans of primary and new lesions in Cohort. **B** (A) The maximum diameter of the primary lesion on baseline and follow-up 2nd MRI scans. (C) The maximum diameter of the largest new lesion appearing on the 2nd MRI scan and its change on the follow-up 3rd MRI scan. (B) The ADC values of the primary lesion on baseline and follow-up 2nd MRI scans. (D) The ADC value of the largest new lesion detected on the second MRI and its change on the subsequent third MRI.

**Table 4.** Baseline MRI characteristics of primary tumors in Cohort B (n=16)

| **Radiological Characteristics** | **CABL-positive group**<br>**(n=15)** | **CABL-negative group**<br>**(n=1)** |
| --- | --- | --- |
| **Lesion Number (n, %)** |  |  |
| single | 6 (40) | 1 (100) |
| multiple | 9 (60) | 0 (0) |
| **BPE\*(n, %)** |  |  |
| < 50% | 12 (80) | 1 (100) |
| ≥50% | 3 (20) | 0 (0) |
| **Patterns of enhancement**<br>**(n, %)** |  |  |
| mass-like | 11 (73.3) |  |
| heterogeneous enhancement | 7 (63.6) | 0 (0) |
| rim enhancement | 4 (36.4) | 0 (0) |
| non-mass-like | 4 (26.7) |  |
| segmental enhancement | 3 (75) | 0 (0) |
| multiregional enhancement | 1 (25) | 1 (100) |
| **Dmax\* /mm** | 40.2 ± 12.0 | 30.9 |
| **Diffusion status (n, %)** |  |  |
| restricted diffusion | 12 (80) | 1 (100) |
| unrestricted diffusion | 3 (20) | 0 (0) |
| **ADC\*(10-3 mm<sup>2</sup>/s)** | 0.976 ± 0.055 | 0.822 |
| **TIC\*(n, %)** |  |  |
| plateau | 12 (80) | 1 (100) |
| wash-out | 3 (20) | 0 (0) |

BPE\*: background parenchymal enhancement; Dmax\*: the maximum diameter; TIC\*: time-intensity curve; ADC\*: apparent diffusion coefficient.

Among the 15 positive patients, CABLs were detected at the second MRI scan in 14 (93.3%) patients, and all of these lesions regressed or disappeared at the third MRI scan. Both mass-like and non-mass-like lesions were markedly enhanced in the early phase of dynamic contrast-enhanced imaging, with signal intensity approaching that of blood vessels, and typically appeared hyperintense relative to the surrounding breast parenchyma on T2-weighted imaging. The radiological features of CABLs are summarized in **Table 5**. Multiple-lesion CABLs were detected in 14 (93.3%), and CABLs of 13 (86.7%) patients involved bilateral breast, while 2 (13.3%) were limited to the contralateral breast. Mass-like lesions were found in 93.3% (14/15), and no mass-like lesions were found in 6.7% (1/15). For mass-like lesions, the median diameter was 12.1 mm (range, 6.8-15.9mm), which decreased to 5.3mm (range, 0-9.0mm) at follow-up (as shown in **Figure 3B**); 7 (50.0%) CABLs had well-defined margins, and 7 (50.0%) had irregular margins; of these, 10 (71.4%) exhibited homogeneous enhancement, and 4 (28.6%) showed heterogeneous enhancement. The non-mass-like lesion measured 14.0 mm and showed linear enhancement. All lesions exhibited unrestricted diffusion on DWI and ADC, with a mean ADC value of (1.847 ± 0.284) ×10□³ mm²/s, which slightly decreased to (1.571 ± 0.313) ×10□³ mm²/s on the next MRI scan (as shown in **Figure 3D**). Notably, the ADC values of primary tumors were significantly lower than those of the new-onset breast lesions (0.976 ± 0.055 vs. 1.847 ± 0.284 ×10□³ mm²/s, *P* < 0.001; **Table 6**). Initially, CABLs in 7 cases (46.7%) showed a wash-out pattern on TIC, 5 cases (33.3%) showed a plateau pattern, and 3 cases (20.0%) showed a wash-in pattern. At follow-up, among 10 evaluable lesions (5 were excluded due to disappearance), CABLs in 6 cases (40.0%) exhibited a plateau pattern, 3 cases (20.0%) exhibited a wash-in pattern, and 1 case (6.7%) exhibited a wash-out pattern (details in **Supplementary Table 4**).

**Table 5.** MRI characteristics of CABLs in Cohort B (n=15)

| **Radiological characteristics** | **CABLs (n=15)** |
| --- | --- |
| **No.of Lesions (n, %)** |  |
| single lesion | 1 (6.7) |
| multifocal lesions | 14 (93.3) |
| **Location (n, %)** |  |
| unilateral (contralateral) | 2 (13.3) |
| bilateral | 13 (86.7) |
| **Onset_time (n, %)** |  |
| 2nd MRI | 14 (93.3) |
| 3rd MRI | 1 (6.7) |
| **Start of recession (n, %)** |  |
| 3rd MRI | 14 (93.3) |
| 4th MRI | 1 (6.7) |
| **Patterns of enhancement (n, %)** |  |
| mass-like | 14 (93.3) |
| homogeneous enhancement | 10 (71.4) |
| heterogeneous enhancement | 4 (28.6) |
| non-mass-like | 1 (6.7) |
| linear enhancement | 1 (100) |
| **Margin of mass-like lesion (n, %)** |  |
| clear | 7 (50.0) |
| irregularity | 7 (50.0) |
| **Dmax\* /mm** |  |
| Mass-like | 12.1 [6.8, 15.9] |
| Non-mass-like | 14.0 |
| **Diffusion_status(n, %)** |  |
| unrestricted diffusion | 15 (100) |
| **ADC\* value /×10-3 mm2/s** | 1.847 ± 0.284 |
| **TIC\*(n, %)** |  |
| wash-in | 3 (20.0) |
| plateau | 5 (33.3) |
| wash-out | 7 (46.7) |
| **Follow_up_Dmax /mm** | 5.3 [0, 9.0] |
| **Follow_up_ADC_value /×10-3 mm2/s** | 1.571 ± 0.313 |
| **Follow_up_TIC(n, %)** |  |
| wash-in | 3 (20.0) |
| plateau | 6 (40.0) |
| wash-out | 1 (6.7) |
| missing | 5 (33.3) |

Dmax\*: the maximum diameter; TIC\*: time-intensity curve; ADC\*: apparent diffusion coefficient.

**Table 6.** The ADC values between primary tumors and CABLs in positive group of Cohort B.

| **Parameters /×10<sup>-3</sup> mm<sup>2</sup>/s** | **Mean±SD** | **P-value** |
| --- | --- | --- |
| **ADC** (Primary lesion) | 0.976 ± 0.055 | *P* <0.001 |
| **ADC** (breast lesion) | 1.847 ± 0.284 |  |

**Figures 4-7** showed the follow-up imaging characteristics and pathological results of three representative CABL-positive cases.

**Figure 4.**
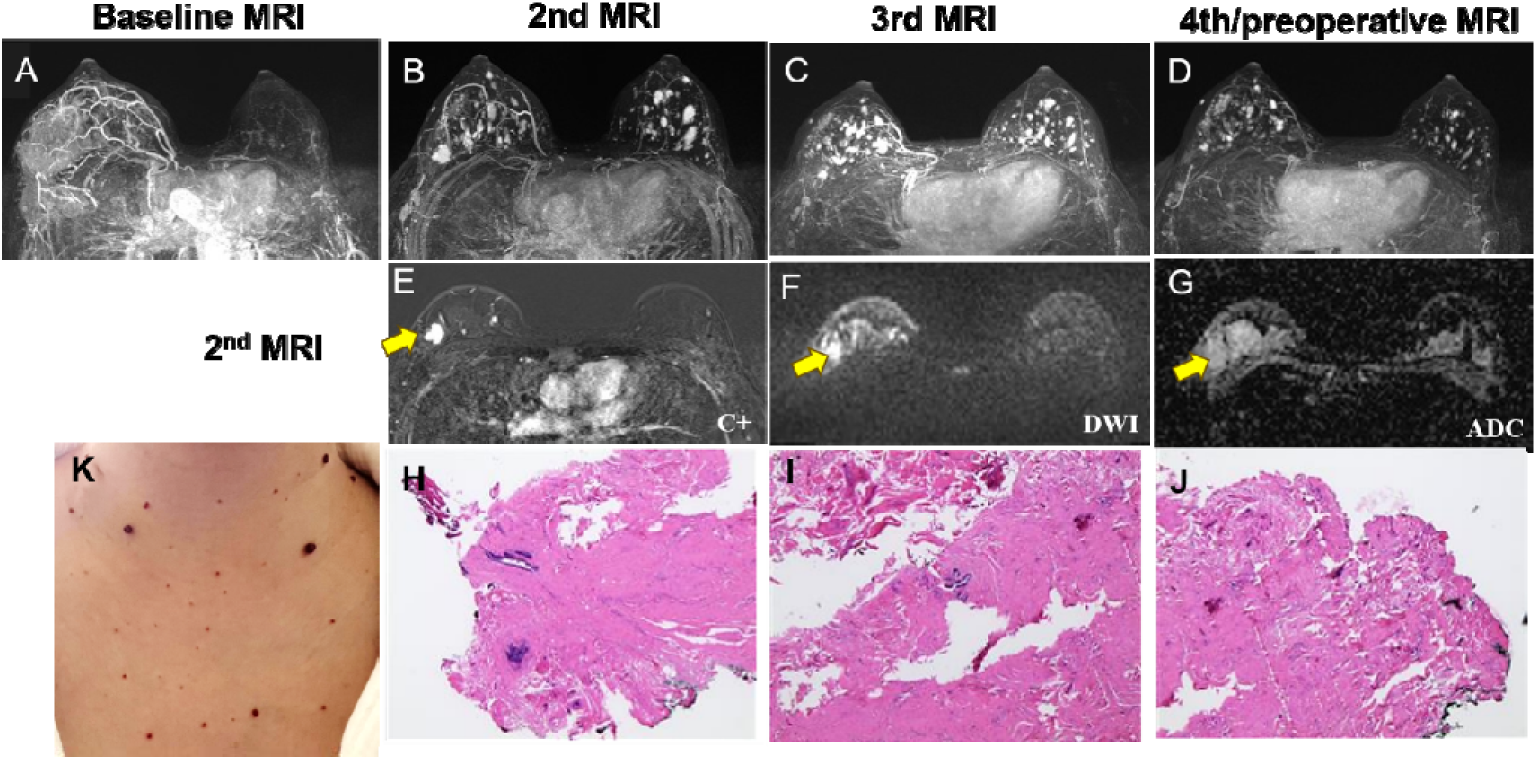
A 30’s years old woman with invasive ductal carcinoma of the right breast; pathology indicates TNBC. (A) A giant lesion with mass-like enhancement in the right breast at baseline MRI. (B) The 2nd MRI review after neoadjuvant therapy suggests centripetal regression of the lesion, with multiple new foci of lesion-like enhancement in both breasts. (C-D) Repeat MRI suggests a slight reduction of the right breast lesion, with further reduction of the new foci of lesion-like enhancement in both breasts. (E-G) The 2nd MRI scan showed regression of the primary lesion and the appearance of multiple new enhancing lesions in both breasts, the largest of which was located in the upper outer quadrant of the right breast; diffusion was unimpeded. (H-J) Biopsy of the new lesions was performed during neoadjuvant therapy; the pathology report indicates breast tissue with lymphocytic infiltration, but no evidence of malignancy was found. (K) Reactive cutaneous capillary endothelial proliferation (RCCEP) was observed in the neck and anterior chest.

**Figure 5.**
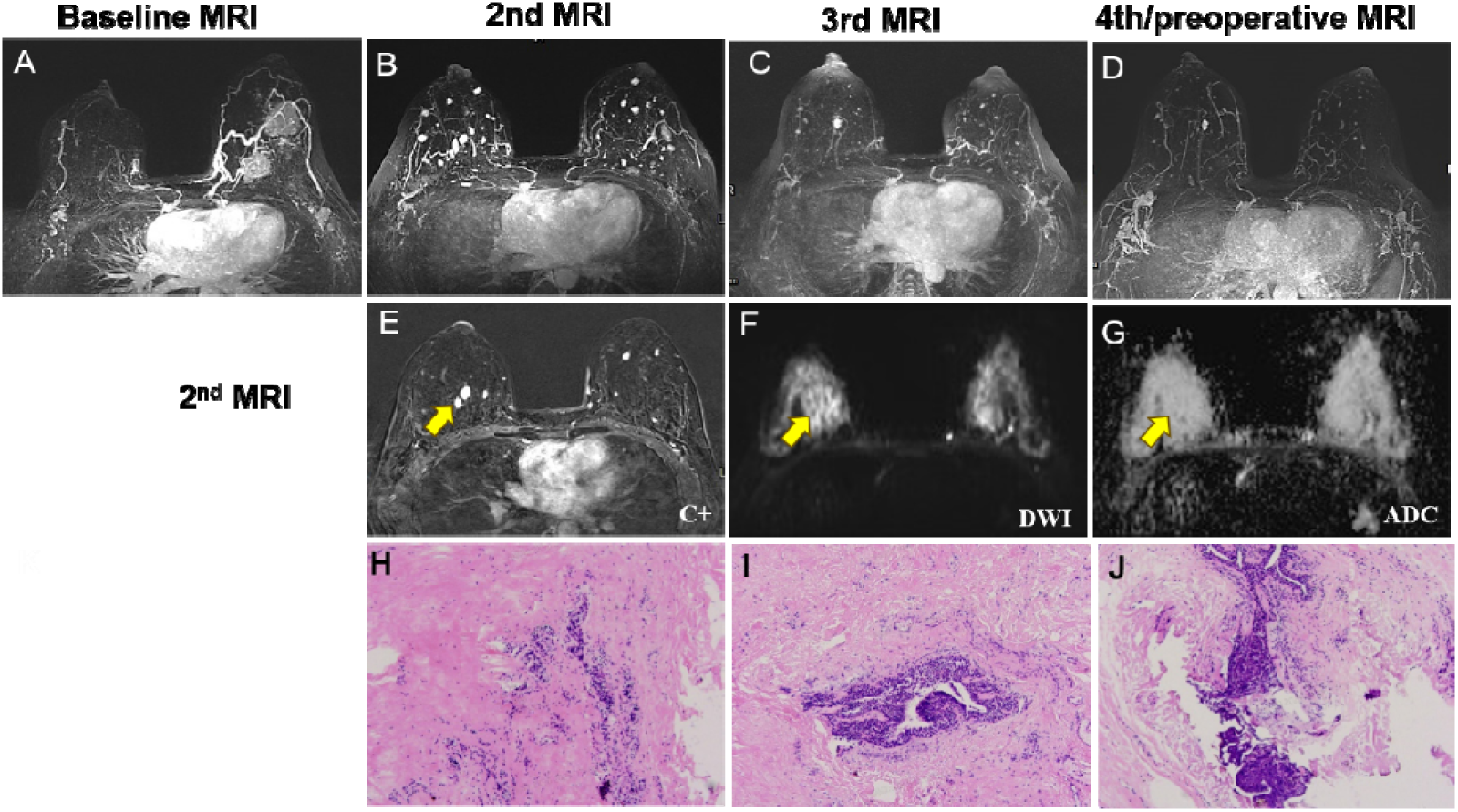
Aged 30’s, invasive ductal carcinoma of the left breast; pathology indicates TNBC. Baseline MRI (A) shows mass-like enhancement in the left breast. (B) The 2nd MRI review after neoadjuvant therapy demonstrates centripetal regression of the lesion and multiple new nodular enhancement foci in both breasts. (C-D) Continued review of the MRI indicates that the primary lesion has achieved imaging complete remission and that the new enhancement foci in both breasts have largely regressed. (E-G) The 2nd MRI scan showed regression of the primary lesion and the appearance of multiple new enhancing lesions in both breasts, the largest of which was located at the 3 o’clock position on the right breast; diffusion was unimpeded. (H-J) Biopsy of the new lesions was performed during neoadjuvant therapy; the pathology report indicates significant proliferation of fibrous tissue in the breast stroma; no clear evidence of cancer was observed.

**Figure 6.**
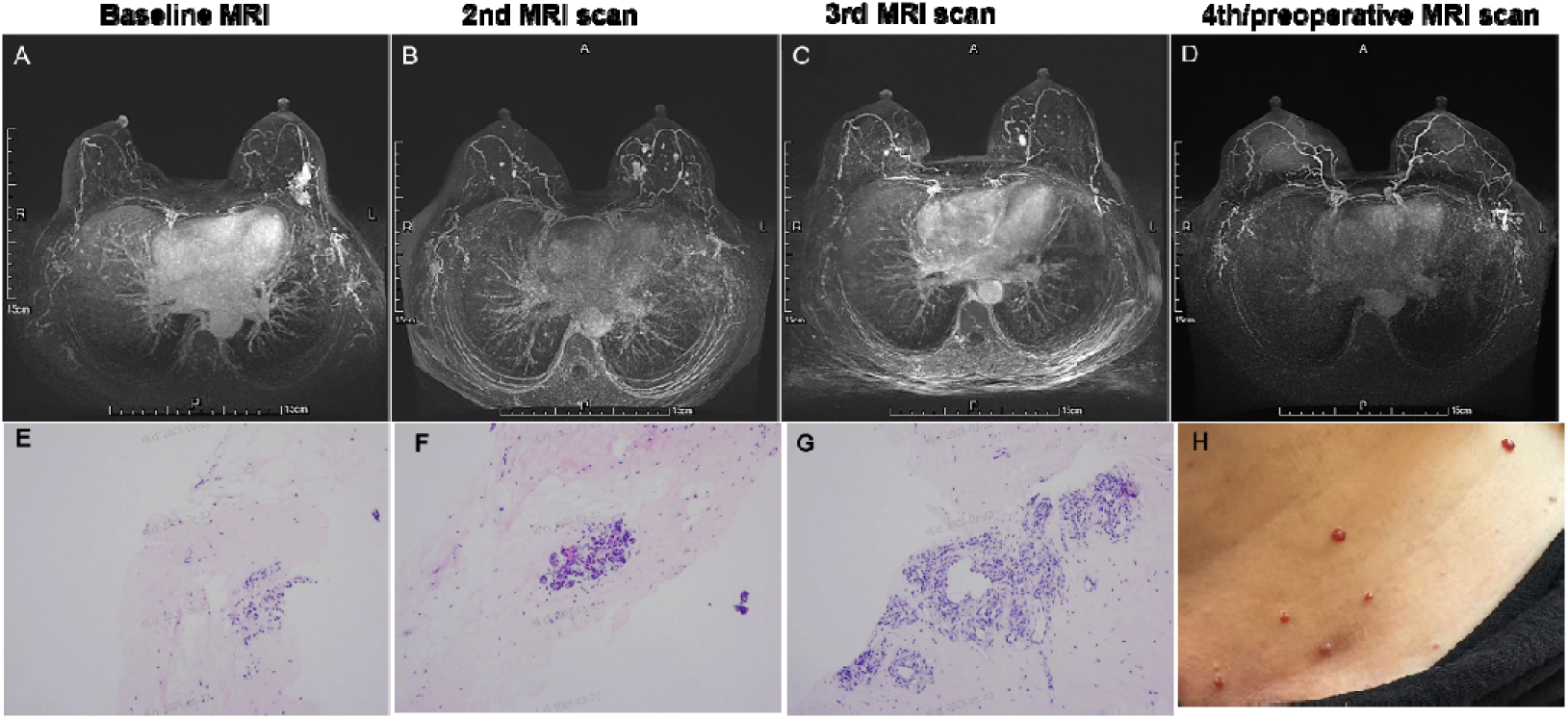
A 50’s years old woman with breast cancer in the left breast; the pathology report indicates Luminal B. Baseline MRI (A) shows a single mass-like enhancement in the left breast. (B) The 2nd MRI review after neoadjuvant therapy demonstrates centripetal regression of the lesion and multiple new nodular enhancement foci in both breasts. (C-D) Continued review of the MRI indicates that the primary lesion has achieved imaging complete remission and that the new enhancement foci in both breasts have largely regressed. (E-F) Biopsy of the new lesions was performed during neoadjuvant therapy; The pathology report indicates a small amount of breast tissue in the right breast, with evidence of adenosis. (H) Reactive cutaneous capillary endothelial proliferation (RCCEP) was observed on the skin of the neck.

**Figure 7.**
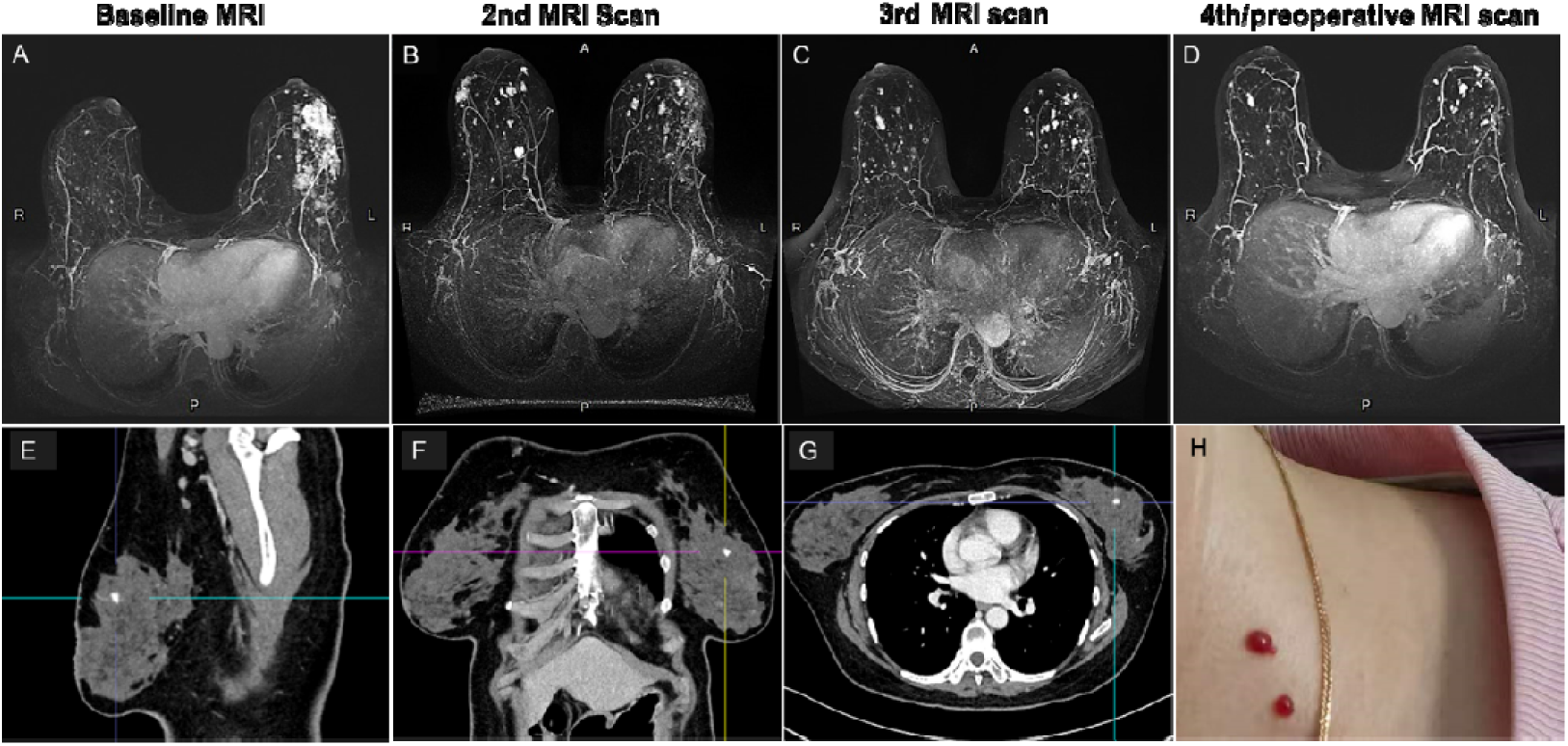
A 40’s years old woman with breast cancer in the left breast; the pathology report indicates Luminal A. (A) Baseline MRI demonstrated segmentally distributed multifocal enhancing lesions in the left breast. (B) Follow-up MRI after 2 cycles of neoadjuvant therapy revealed nonconcentric regression of the left breast lesion, with new bilateral nodular enhancing foci. (C) The third MRI showed further regression of the left breast lesion and interval shrinkage of bilateral nodular foci. (D) Preoperative MRI confirmed continued regression of all lesions. (E-G) The fourth follow-up contrast-enhanced chest CT scan revealed a small enhanced lesion in the left breast; (H) Reactive cutaneous capillary endothelial proliferation (RCCEP) was observed on the skin of the neck.

The sole patient who did not develop CABL was a 60s years old postmenopausal woman with a family history of breast cancer. The breast cancer was a stage III, high-grade, luminal B invasive ductal carcinoma harboring a BRCA2 mutation, with lymph node metastasis. She achieved pCR after neoadjuvant therapy. Due to the small sample size (n=1 in the CABL-negative group), no statistical comparisons were performed between the two subgroups. On baseline MRI, the breast cancer presented as a single non-mass-like enhancement with multiregional distribution, restricted diffusion, and minimal to mild BPE (BPE<50%), and showed a plateau pattern on TIC.

## Discussion

IrAEs can affect nearly every organ system, most commonly the skin, gastrointestinal tract, liver, lung, and endocrine organs. However, its involvement of the breast—whether as a primary or secondary site—has been rarely reported^[5]^. This phenomenon may be explained by the breast being neither an immune-privileged site nor a site of intense immune surveillance, which renders breast irAEs rare and prone to misdiagnosis. In this dual-cohort retrospective study, we systematically characterized CABLs in both non-breast cancer and breast cancer patients. In our study, 34.4% (62/180) of patients in Cohort A and 93.8% (15/16) of patients in Cohort B developed CABLs during camrelizumab treatment, and we considered them to be irAEs rather than true disease progression. These lesions were highly consistent with irAEs in onset timing and clinical behavior. First, the temporal onset (mostly 2-3 cycles, approximately 2–3 months) falls within the typical irAE window of 1.4–6.4 months after ICIs initiation^[3, 6]^. Second, the self-limited course (40.3% disappearance, 46.8% shrinkage) aligns with the known behavior of reactive lesions, such as RCCEP, which spontaneously resolve after drug discontinuation^[4, 7]^. Third, some patients in our cohort simultaneously or sequentially experienced other irAEs, including RCCEP and rash.

Across different tumor types, these lesions exhibited consistent radiological features: multifocal, mass-like, well-circumscribed, homogeneously enhancing lesions with unrestricted diffusion. Premenopausal patients were more susceptible, and the lesions typically appeared within 2–3 cycles after treatment initiation (or by the second MRI scan), subsequently disappearing or shrinking in the majority of cases. Notably, CABL-positive patients in both cohorts showed a trend toward better treatment response (higher ORR in Cohort A and pCR rate of 53.3% in Cohort B), suggesting favorable prognostic implications. Reports of breast lesions arising after this immunotherapy remain scarce. By detailing their imaging characteristics and pattern of evolution, this study offers critical information that may help clinicians avoid misdiagnosis and missed diagnosis.

On CT and MR imaging, these CABLs were characterized by multifocal, mass-like, well-circumscribed, homogeneously enhancing lesions with unrestricted diffusion. There was a difference in lesions between the two groups. Unilateral involvement was observed in 66.1% (41/62) of cases in Cohort A and bilateral involvement in 86.7% (13/15) of cases in Cohort B. The median lesion diameter was smaller in Cohort A than in Cohort B (7 mm vs 14.1 mm). The difference in distribution and diameter was likely due to the inferior soft-tissue resolution of CT relative to MRI, compounded by the supine position used for chest CT, which compresses and deforms these breast lesions, even rendering them inconspicuous. Moreover, the dynamic evolution of these lesions also aligns with pseudoprogression (PsP). PsP<u>—</u>a kind of atypical response patterns following immunotherapy<u>—</u>manifests as temporary tumor enlargement or new lesion development (with a total tumor burden of ≥20%) without significant clinical deterioration, followed by delayed regression or disappearance with continued immunotherapy ^[8]^. In Cohort B, the appearance of CABLs was not linked to rising overall tumor burden; the mean diameter of primary tumors decreased from 40.2±12.0 mm at baseline to 20.2±14.1 mm at follow-up, while the mean ADC value increased from 0.976 ± 0.055 ×10LJ³ mm²/s to 1.847 ± 0.284 ×10LJ³ mm²/s. All patients exhibited systemic disease control during nodule stabilization or regression.

Histopathological findings of several CABLs in our study further confirmed their inflammatory nature. In Cohort B, CABLs of four cases underwent biopsy or surgery, showing lymphocytic infiltration and fibrous tissue proliferation with no evidence of malignancy. Taken together, these clinical, radiological, and pathological features prompted us to explore the possible pathogenesis of such breast lesions. **Figure 8 shows the Biological Mechanisms Maps**.

**Figure 8.**
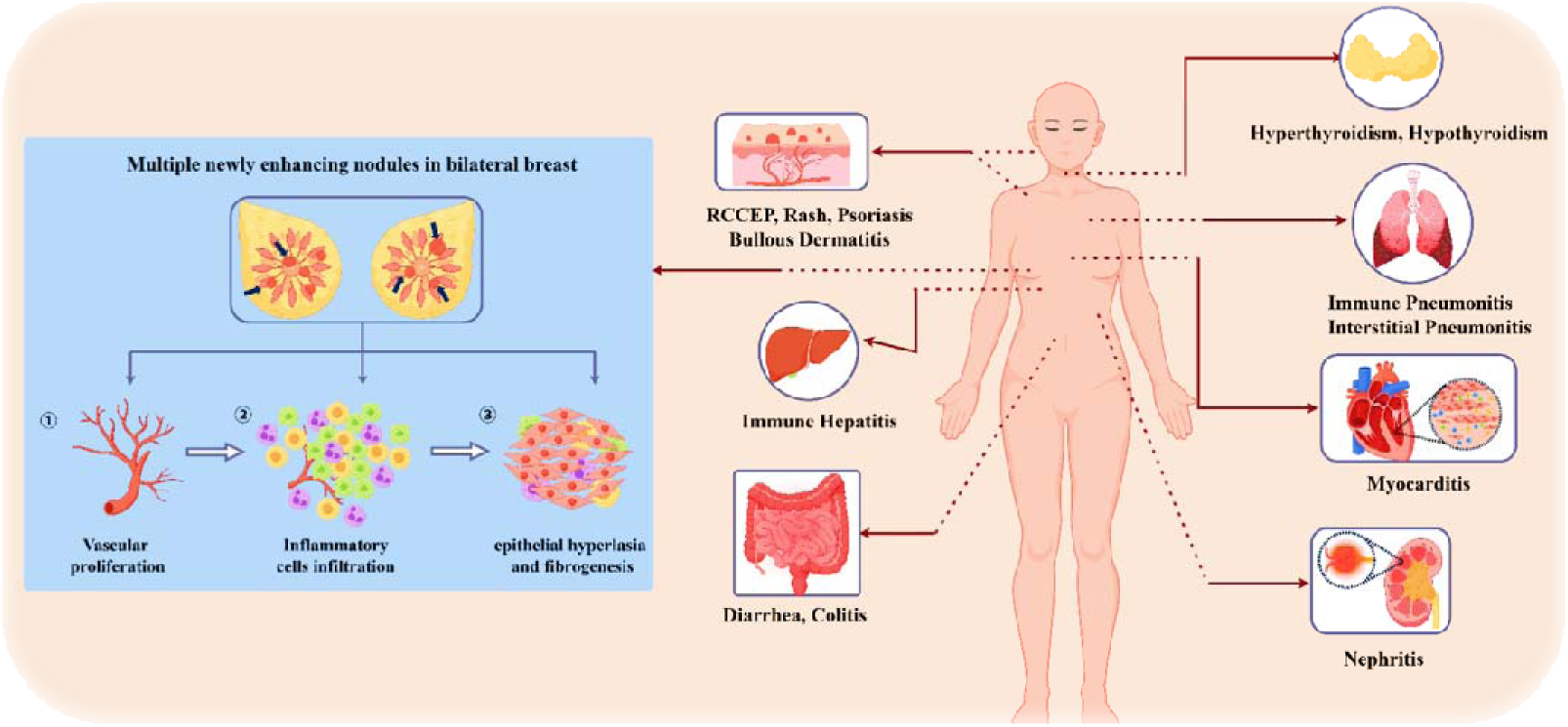
Biological Mechanisms Maps. Camrelizumab’s systemic adverse reactions and possible biological mechanisms related to the breast

Common adverse reactions to Carrelizumab affect major organs throughout the body. Skin: reactive telangiectasia, rash, pruritus; Thyroid: Hypothyroidism, hyperthyroidism; Lungs: Immune-mediated pneumonia, interstitial pneumonia; Liver: Elevated liver enzymes, immune-mediated hepatitis; Gastrointestinal tract: Diarrhoea, colitis, nausea and vomiting; Kidneys: Renal impairment, immune-mediated nephritis; Heart: Myocarditis, arrhythmias, etc. This study focuses on adverse reactions in the breast, manifested as new bilateral enhanced lesions; the possible mechanisms underlying this are as follows: ⍰ Angiogenic Modulation: Somatic gene mutations, combined with PARP inhibitor-induced dysregulation of angiogenic pathways, may promote aberrant neovascularization, potentially manifesting as de novo nodule formation on imaging studies. ⍰ Immune-Mediated Pseudomorphology: Treatment-related inflammatory cell infiltration within tumor microenvironments could induce transient lesion expansion through immune cell margination and peritumoral activation. ⍰ Stromal Remodeling: Persistent inflammatory microenvironments may drive pathological fibroblast activation, potentially leading to either reactive desmoplasia or discrete fibrotic tissue formation.

### Immune-Mediated Pseudoprogression

Previous research^[9]^ has suggested that PsP mechanisms may involve complex microenvironmental changes: (1) continued tumor progression until effective antitumoral immune response activation; (2) immune-related inflammation or edema resulting from lymphocyte infiltration; (3) local vascular rupture, hemorrhage, or accumulation of tumor necrosis products^[9]^. The pathological evidence from our study confirmed that inflammatory cell infiltration and fibroblast proliferation were responsible for the formation of these breast lesions.

### Camrelizumab-Induced Capillary Proliferation

As a distinctive irAE uniquely associated with camrelizumab^[4]^, RCCEP exhibited the temporal similarity and shared angiomorphic features with these breast lesions, suggesting a common pathogenesis. Camrelizumab’s unique pharmacological properties also played an important role. Camrelizumab aberrantly binds to vascular endothelial growth factor receptor 2 (VEGFR2) on capillary endothelial cells, directly stimulating proliferation^[10]^. Concurrently, it may trigger excessive immune activation, disrupting the local balance between pro- and anti-angiogenic factors, thereby promoting benign endothelial proliferation^[11]^. It also activates CD4□ T cells, increasing Th2 cytokine IL-4, which promotes differentiation of CD163□ M2 macrophages and VEGF-A release, driving benign endothelial hyperplasia^[4]^. Nevertheless, typical vascular proliferation was not observed on the pathology of these breast nodules, possibly owing to the timing of biopsy or surgical excision.

### BRCA Status and Pre-Existing Angiogenic Priming

Converging evidence indicates that BRCA1/2-deficient breast cancers exhibit upregulated angiogenesis, with elevated levels of VEGF, Ang-1, and Ang-2 ^[12–14]^. In our breast cancer cohort, the high incidence of lesions (93.8%) paralleled the high frequency of *BRCA1/2* mutations (40% and 33.3%, respectively). Previous studies have shown that *BRCA1*-deficient breast cancers exhibit upregulated angiogenesis, with elevated VEGF, Ang-1, and Ang-2 levels, and that BRCA1 normally represses VEGF transcription via the CtIP/ZBRK1 complex^[12–14]^. This pre-existing angiogenic vulnerability may lower the threshold for camrelizumab-induced capillary proliferation, explaining the markedly higher lesion incidence in breast cancer patients compared to those with other malignancies. Thus, the formation of CABLs appears to result from the interplay of camrelizumab-specific angioproliferation, inflammatory remodeling, and pre-existing angiogenic vulnerability associated with BRCA mutations.

Our findings align with those of a recent study by Ma et al^[15]^, who reported that 58.7% of triple-negative breast cancer (TNBC) patients receiving camrelizumab-based neoadjuvant therapy developed new enhancing breast lesions on MRI, which also regressed with continued treatment. However, our work extends that report in two important respects. First, we demonstrate that these lesions are not restricted to TNBC: the majority of our CABL-positive breast cancer patients were luminal B subtype (46.7%), and similar lesions occurred in non-breast cancer patients, indicating a broader phenomenon. Second, whereas Ma et al^[15]^ proposed that these lesions represent camrelizumab-induced benign vascular proliferation within the breast, our pathological findings did not show direct evidence of intralesional microvascular proliferation. We therefore consider these lesions to result from a multifactorial process, rather than a purely angioproliferative phenomenon. Given the discrepancies between the two studies, further investigations with standardized protocols and larger cohorts are warranted to fully characterize the nature and pathogenesis of these lesions.

A growing body of evidence links mild to moderate irAEs development with improved ICI efficacy ^[16–20]^. Consistently, patients experiencing PsP have been reported to exhibit shorter response duration than those with typical responses, yet significantly better survival outcomes than those with true disease progression^[21, 22]^. In Cohort A, CABL□positive patients showed a higher ORR (58.1% vs. 50.0%) and a lower proportion of PD (3.2% vs. 17.8%); in Cohort B, 53.3% of CABL□positive patients achieved pCR. This pCR rate substantially exceeds the 30–40% typically observed with standard neoadjuvant chemotherapy and the 18% reported for HR+/HER2□negative breast cancer ^[23–27]^. The association was further supported by the statistically significant difference in the distribution of best overall response between CABL□positive and CABL□negative groups in Cohort A (P=0.031). Similar to RCCEP, which has been shown to correlate with higher ORR, longer PFS, and OS across multiple cancers^[4, 18, 28, 29]^, the development of these breast lesions may serve as a clinical biomarker of robust immune activation and enhanced antitumor immunity. Therefore, their presence should not prompt treatment discontinuation but rather be recognized as a potential positive signal.

In cancer patients, CABLs, particularly mass-like lesions, are typically suspected to represent metastases or residual disease. This underscores the need to include irAEs in the differential diagnosis when solitary breast nodules develop during immunotherapy. Although most breast lesions in our study exhibited benign properties, some proved challenging to distinguish from true progressive disease, with 46.7% (7/15) lesions showing irregular margins and 46.7% (7/15) lesions displaying a washout pattern on TIC. In this context, ADC values may aid in the differentiation by reflecting microstructural changes. In our series, ADC values of these lesions consistently exceeded the accepted threshold for malignancy (typically <1.00–1.23 × 10□³ mm²/s) ^[30, 31]^, thereby further bolstering diagnostic confidence on MRI. According to iRECIST guidelines, any new lesion should initially be classified as “unconfirmed progressive disease” (iUPD), requiring repeat imaging after 4–8 weeks to confirm progression^[32]^. Our findings support this approach: the majority of lesions appeared at the second MRI scan (median 3 cycles) and regressed by the third scan. For clinically stable patients, we advocate a **watch-and-wait strategy** with short-interval MRI (e.g., 4–6 weeks) before considering biopsy or changing therapy. This strategy aligns with established practice for PsP and can safely avoid unnecessary invasive procedures and premature treatment cessation e^[3, 30, 32, 33]^. However, any lesion that exhibits persistent growth or fails to regress within the expected timeframe should undergo histopathological clarification.

## Limitations

Several limitations should be acknowledged. First, the sample size of Cohort B is small, and the CABL-negative group consisted of only one patient, precluding statistical comparisons. Second, only four lesions were confirmed by histopathology; most relied on follow-up MRI, lacking tissue diagnosis. Future prospective studies with larger cohorts, mandatory biopsy of lesions, and correlative biomarker analyses (e.g., VEGFR2 expression, T-cell infiltration) are warranted to validate our findings and elucidate the underlying biology.

## Conclusions

To our knowledge, this is the first report to describe such lesions across diverse tumor types and to integrate imaging, pathological, and clinical outcome data. CABLs are predominantly benign reactive changes associated with immunotherapy. Their radiological and temporal features, together with pathological confirmation in a subset, support classification as an irAE. Premenopausal status and underlying BRCA-related angiogenic susceptibility may increase risk. The development of these lesions appears to signal favorable treatment outcomes, potentially serving as a clinical biomarker. A watch-and-wait approach with imaging follow-up is recommended to avoid unnecessary interventions, while maintaining vigilance for atypical behavior.

## Disclosure paragraph

## Data Availability

All data generated in this study are included in the manuscript.

## Acknowledgements

We thank all the members of the institution for their help in discussions and for providing technical assistance. We thank Home for Researchers editorial team (www.home-for-researchers.com) for table and language editing services.

## Statistics and Biometry

No complex statistical methods were necessary for this paper.

## Informed Consent

Written informed consent was waived by the Institutional Review Board.

## Study subjects or cohorts overlap

The study subjects or cohorts have not been previously reported.

## Methodology

– retrospective
– observational study
– performed at one institution

