## Supplementary table for "A Radiologic Masquerade: Camrelizumab-Associated Breast Lesions That Mimic Progression"

| **Supplementary Table 1. Scanning parameters for CT examination** | | | | |
| --- | --- | --- | --- | --- |
| **CT scanners** | **Tube current (mA)** | **Tube voltage (kV)** | **Slice thickness (mm)** | **Pixel spacing** |
| Philips IQon Spectral; Canon Aquilion Prime; Siemens Somatom Force; GE Discovery CT750 HD | 143-250 | 120 | 1 | 0.609-0.797 |

| **Supplementary Table 2. Scanning parameters for MRI sequences** | | | | | | | |
| --- | --- | --- | --- | --- | --- | --- | --- |
| **Scanning parameters** | **TR/TE** | **PD** | **FOV** | **MS** | **SL** | **SR** | **ST** |
| T1-DCE | 4.32/1.57 | 446 | 380×380 | 448×448 | 144 | 0.848×0.848 | 1.0 |
| FS-T2WI | 4330/61 | 319 | 380×380 | 320×320 | 38-42 | 1.188×1.188 | 3.0 |
| ADC map | 6300/74 | 2083 | 380×380 | 160×160 | 24-32 | 2.375×2.375 | 4.0 |
| *TR/TE,repeat time/echo time (ms); PD,Pixel bandwidth;FOV, field of view (mm); MS, Matrix size;SL,Slicer layer;SR, Spatial resolution (mm2); ST,Slicer thickness (mm); T1-DCE, T1-weighted dynamic contrast-enhancement imaging; FS-T2WI, Fat suppression T2-weighted imaging; ADC map,apparent diffusion coefficient map.* | | | | | | | |

| **Supplementary Table 3. The qualitative and quantitative radiological features of primary tumors in Cohort B (n=16)** | | | | | | | | | | | |
| --- | --- | --- | --- | --- | --- | --- | --- | --- | --- | --- | --- |
|  | **Baseline** | | | | | | | |  | **The 2nd MRI scan** | |
| **Patient ID** | **Number** | **Intratumoral hyperintensity in T2WI** | **BPE^*^** | **Patterns of enhancement** | | **Diffusion status** | **TIC^*^** | **D_max_^*^ /mm** | **ADC value /×10^-3^ mm^2^/s** | **D_max_^*^ /mm** | **ADC value /×10^-3^ mm^2^/s** |
| 1 | single | absence | ＜50% | mass-like | rim enhancement | restricted | plateau | 43.0 | 1.176 | 21.0 | 1.067 |
| 2 | multiple | presence | ≥50% | mass-like | rim enhancement | restricted | plateau | 33.0 | 0.773 | 21.0 | 1.060 |
| 3 | multiple | absence | ＜50% | non-mass-like | segmental enhancement | unrestricted | plateau | 59.0 | 0.828 | 37.5 | 1.225 |
| 4 | multiple | absence | ≥50% | non-mass-like | multiregional enhancement | unrestricted | wash-out | 45.0 | 0.994 | 25.5 | 1.131 |
| 5 | single | absence | ＜50% | mass-like | heterogeneous enhancement | restricted | plateau | 40.5 | 0.938 | 33.0 | 1.091 |
| 6 | single | presence | ＜50% | mass-like | heterogeneous enhancement | restricted | plateau | 52.0 | 0.791 | 22.0 | 0.978 |
| 7 | multiple | presence | ＜50% | non-mass-like | segmental enhancement | restricted | plateau | 51.5 | 1.070 | 50.0 | 1.858 |
| 8 | multiple | absence | ＜50% | mass-like | rim enhancement | restricted | wash-out | 15.5 | 0.822 | 0.0 | 1.336 |
| 9 | single | presence | ＜50% | mass-like | heterogeneous enhancement | restricted | plateau | 29.5 | 0.768 | 21.0 | 1.130 |
| 10 | multiple | absence | ＜50% | non-mass-like | segmental enhancement | restricted | plateau | 31.0 | 0.946 | 0.0 | 1.251 |
| 11 | multiple | absence | ＜50% | mass-like | heterogeneous enhancement | restricted | plateau | 51.0 | 0.817 | 17.0 | 1.492 |
| 12 | single | absence | ≥50% | mass-like | heterogeneous enhancement | restricted | wash-out | 31.5 | 0.958 | 26.5 | 1.153 |
| 13 | multiple | presence | ＜50% | mass-like | rim enhancement | restricted | plateau | 34.0 | 1.265 | 0.0 | 1.364 |
| 14 | multiple | absence | ＜50% | mass-like | heterogeneous enhancement | restricted | plateau | 32.0 | 0.958 | 10.0 | 0.898 |
| 15 | single | absence | ＜50% | mass-like | heterogeneous enhancement | unrestricted | plateau | 54.0 | 1.543 | 18.0 | 1.109 |
| 16 | single | presence | ＜50% | non-mass-like | multiregional enhancement | restricted | plateau | 30.9 | 0.822 | 5.4 | 1.440 |
| D_max_^*^: the maximum diameter; BPE^*^: background parenchymal enhancement; TIC^*^: time-intensity curve. | | | | | |  |  |  |  |  |  |

| **Supplementary Table 4. The qualitative and quantitative radiological features of Cohort B's CABLs (n=15)** | | | | | | | | | | | | | | | | |
| --- | --- | --- | --- | --- | --- | --- | --- | --- | --- | --- | --- | --- | --- | --- | --- | --- |
|  |  |  |  |  |  |  |  |  |  | **The 2nd MRI scan** | |  | **The 3rd MRI scan** | | **The 4th MRI scan** | |
| **Patient ID** | **The initial emergence time** | **Start of recession** | **Location** | **Number** | **Margin** | **Patterns of enhancement** | | **Diffusion status** | **TIC^*^** | **D_max_^*^ /mm** | **ADC value /×10^-3^ mm^2^/s** | **Follow up TIC^*^** | **D_max_^*^ /mm** | **ADC value /×10^-3^ mm^2^/s** | **D_max_^*^ /mm** | **ADC value /×10^-3^ mm^2^/s** |
| 1 | 2nd^*^ | 3rd^*^ | Bilateral | multiple | clear | mass-like | heterogeneous enhancement | unrestricted | wash-in | 10.0 | 1.154 | - | 0.0 | 1.008 | - | - |
| 2 | 2nd^*^ | 3rd^*^ | Bilateral | multiple | clear | mass-like | homogeneous enhancement | unrestricted | wash-out | 14.5 | 1.665 | wash-out | 8.3 | 1.420 | 7.6 | 1.356 |
| 3 | 2nd^*^ | 3rd^*^ | Bilateral | multiple | irregularity | mass-like | heterogeneous enhancement | unrestricted | plateau | 15.5 | 1.896 | plateau | 8.0 | 1.587 | 6.5 | 1.631 |
| 4 | 2nd^*^ | 3rd^*^ | Unilateral | single | clear | mass-like | homogeneous enhancement | unrestricted | plateau | 6.8 | 1.789 | wash-in | 3.0 | 1.404 | - | - |
| 5 | 2nd^*^ | 3rd^*^ | Bilateral | multiple | clear | mass-like | homogeneous enhancement | unrestricted | wash-in | 7.0 | 1.809 | - | 0.0 | 1.709 | 0.0 | 1.922 |
| 6 | 3rd^*^ | 4th^*^ | Bilateral | multiple | clear | mass-like | homogeneous enhancement | unrestricted | - | - | - | wash-out | 8.5 | 1.914 | 7.0 | 1.708 |
| 7 | 2nd^*^ | 3rd^*^ | Bilateral | multiple | clear | mass-like | homogeneous enhancement | unrestricted | plateau | 9.0 | 2.086 | wash-in | 4.0 | 2.067 | 4.0 | 2.035 |
| 8 | 2nd^*^ | 3rd^*^ | Bilateral | multiple | clear | non-mass-like | linear enhancement | unrestricted | wash-out | 14.0 | 1.544 | plateau | 6.0 | 1.381 | 0.0 | 1.391 |
| 9 | 2nd^*^ | 3rd^*^ | Bilateral | multiple | clear | mass-like | homogeneous enhancement | unrestricted | plateau | 9.5 | 1.902 | plateau | 5.5 | 1.900 | 4.5 | 1.708 |
| 10 | 2nd^*^ | 3rd^*^ | Bilateral | multiple | irregularity | mass-like | heterogeneous enhancement | unrestricted | plateau | 14.2 | 1.939 | plateau | 9.0 | 1.898 | 8.0 | 1.696 |
| 11 | 2nd^*^ | 3rd^*^ | Bilateral | multiple | irregularity | mass-like | homogeneous enhancement | unrestricted | wash-out | 15.0 | 1.717 | plateau | 9.0 | 1.420 | 7.0 | 1.602 |
| 12 | 2nd^*^ | 3rd^*^ | Unilateral | multiple | irregularity | mass-like | heterogeneous enhancement | unrestricted | wash-out | 15.0 | 2.287 | wash-in | 5.0 | 1.256 | 4.5 | 1.404 |
| 13 | 2nd^*^ | 3rd^*^ | Bilateral | multiple | irregularity | mass-like | homogeneous enhancement | unrestricted | wash-out | 10.0 | 1.980 | - | 0.0 | 1.293 | 0.0 | 2.166 |
| 14 | 2nd^*^ | 3rd^*^ | Bilateral | multiple | irregularity | mass-like | homogeneous enhancement | unrestricted | wash-in | 15.9 | 2.221 | plateau | 8.4 | 2.005 | 8.4 | 2.051 |
| 15 | 2nd^*^ | 3rd^*^ | Bilateral | multiple | irregularity | mass-like | homogeneous enhancement | unrestricted | wash-out | 15.0 | 1.862 | - | 0.0 | 1.644 | 0.0 | 1.927 |
| D_max_^*^: the maximum diameter; TIC^*^: time-intensity curve; 2nd^*^: the second MRI scan; 3rd^*^: the third MRI scan; 4th^*^: the fourth MRI scan. | | | | | | | | | | | |  |  |  |  |  |
